# Renal Outcomes in Survivors of Neonatal and Pediatric Renal Vein Thrombosis

**DOI:** 10.64898/2026.08.25.26361338

**Authors:** Mayra A. Oseguera, Lena S. Bercz, Joseph R. Stanek, Myda Khalid, Bryce A. Kerlin

**Author notes:** Corresponding Author: Bryce A. Kerlin, MD, Center for Clinical & Translational Research, The Abigail Wexner Research Institute at Nationwide Children’s 700 Children’s Dr., W302, Columbus, OH 43205, USA.

## Abstract

**Introduction:** Pediatric renal vein thrombosis is a rare but well-recognized form of venous thromboembolism. While long-term renal outcomes of neonatal cases are well-described, they are relatively unknown in cases affecting older children. Moreover, the ability of anticoagulation treatment to prevent these outcomes remains unknown. The objective of this study was to assess adverse long-term renal outcomes and determine if anticoagulation reduced their likelihood.

**Methods:** Administrative data analysis utilizing the Pediatric Health Information System database. Renal vein thrombosis occurring in patients under 18 years were assessed. Cases involving “tumor thrombus” were excluded to focus the analysis only on thrombotic disease. Demographics, co-morbid conditions, anticoagulant therapies, and renal outcomes were assessed over a 9-year period. In sub-analyses, neonatal (≤ 28 days) and non-neonatal renal vein thromboses were assessed to determine how their characteristics may differ.

**Results:** 383 eligible renal vein thrombosis cases with 796 patient-years of follow-up were identified for analysis. 48.8% of the cases occurred in neonates. 25.3% of the cases occurred in children with pre-existing complex chronic conditions and 9.1% were associated with the onset of nephrotic syndrome. Mortality followed 15.1% of the cases, but causality cannot be assigned from administrative data. Most (80.2%) of the cases were treated with anticoagulation. Acute kidney injury occurred in 24% of cases, chronic kidney disease developed in 22.2%, hypertension in 26.9%, and proteinuria in 3.1%. Anticoagulation did not have a discernable effect on the likelihood of these long-term renal outcomes.

**Conclusion:** Acute kidney injury, chronic kidney disease, and hypertension are prevalent in survivors of childhood renal vein thrombosis. Anticoagulation does not appear to reduce the incidence of long-term renal outcomes, but the low percentage of non-anticoagulated patients suggests treatment bias. Long-term kidney health surveillance is warranted in pediatric renal vein thrombosis survivors.

## INTRODUCTION

Pediatric renal vein thrombosis (RVT) is a rare but well-recognized form of venous thrombosis that develops in the major renal veins or its tributaries.^1,2^ RVT has been reported to be most common in neonates, accounting for up to 70% of cases, and is the most common form of neonatal thrombosis not associated with central venous catheterization.^3–5^ Although the incidence is uncertain, it is estimated to be around 2 to 5 cases per 100,000 live births.^1,3,4,6^ RVT can also occur in older children, albeit less commonly.^2,5^ Neonatal RVT presents with significant long-term morbidity, including elevated risks of hypertension, chronic kidney disease (CKD), and end stage kidney disease (ESKD).^1,3,6^

The optimal management of pediatric RVT remains unclear.^2^ Current guidelines recommend anticoagulation therapy in neonates with confirmed RVT, particularly in cases of bilateral involvement or thrombus extension into the inferior vena cava, given the risk for bilateral kidney injury in this setting.^3,7–9^ However, there is little prospective evidence to support this approach and, therefore, the guideline recommendation is conditional based on very low certainty in the available evidence about effects.^3,7,10^ Furthermore, because data for renal outcomes following RVT in older children and adolescents are scant, the use of anticoagulation remains controversial and ill-defined.

The objectives of this study were to define the epidemiology of and risk factors for RVT in the neonatal and pediatric populations and to evaluate the efficacy of anticoagulation to prevent detrimental long-term renal outcomes.

## METHODS

### Data Source

Data for this study were obtained from the Pediatric Health Information System (PHIS) database. PHIS is a large administrative database which includes data from inpatient, ambulatory, and emergency department observation-level encounters from 50 of the largest tertiary care pediatric hospitals across the United States. Through an affiliation with the Children’s Hospital Association (CHA) (Lenexa, KS), data is routinely checked for data quality and reliability by both the included hospitals and CHA. Data is de-identified, and patients are given a unique identifier which allows for tracking patient encounters over time. PHIS data includes information on patient demographics, discharge diagnoses, procedures, billing codes for resource utilization (i.e., labs, medications, imaging, supplies, etc.), and healthcare costs. De-identified data is considered exempt by the Nationwide Children’s Hospital Institutional Review Board.

### Study Population and Definitions

PHIS patients aged 0 to 18 years with a diagnosis of RVT between October 2015 through December 2023 were identified (**Fig. 1**). Patients with an RVT diagnosis were determined using the International Classification of Diseases, 10^th^ Revision (ICD-10; **Table S1**). Patients with an RVT were excluded if they were diagnosed with a type of cancer that is associated with “tumor thrombus” (**Table S2**). Following RVT diagnosis, all subsequent inpatient and outpatient encounters were included to assess renal outcomes. ICD-10 codes were used to identify patients who developed kidney disease (i.e., acute kidney injury (AKI), CKD, end-stage kidney disease (ESKD), hypertension, proteinuria, or hematuria (**Table S3**)). Pre-existing kidney disease (i.e., CKD, hypertension, proteinuria, or hematuria) was considered as a risk factor for RVT occurrence, but patients with pre-existing kidney disease were excluded from the analysis of how RVT impacts future kidney disease risk (i.e., after RVT). Patients were included independent of any previous history of anticoagulation treatment.

**Figure 1:**
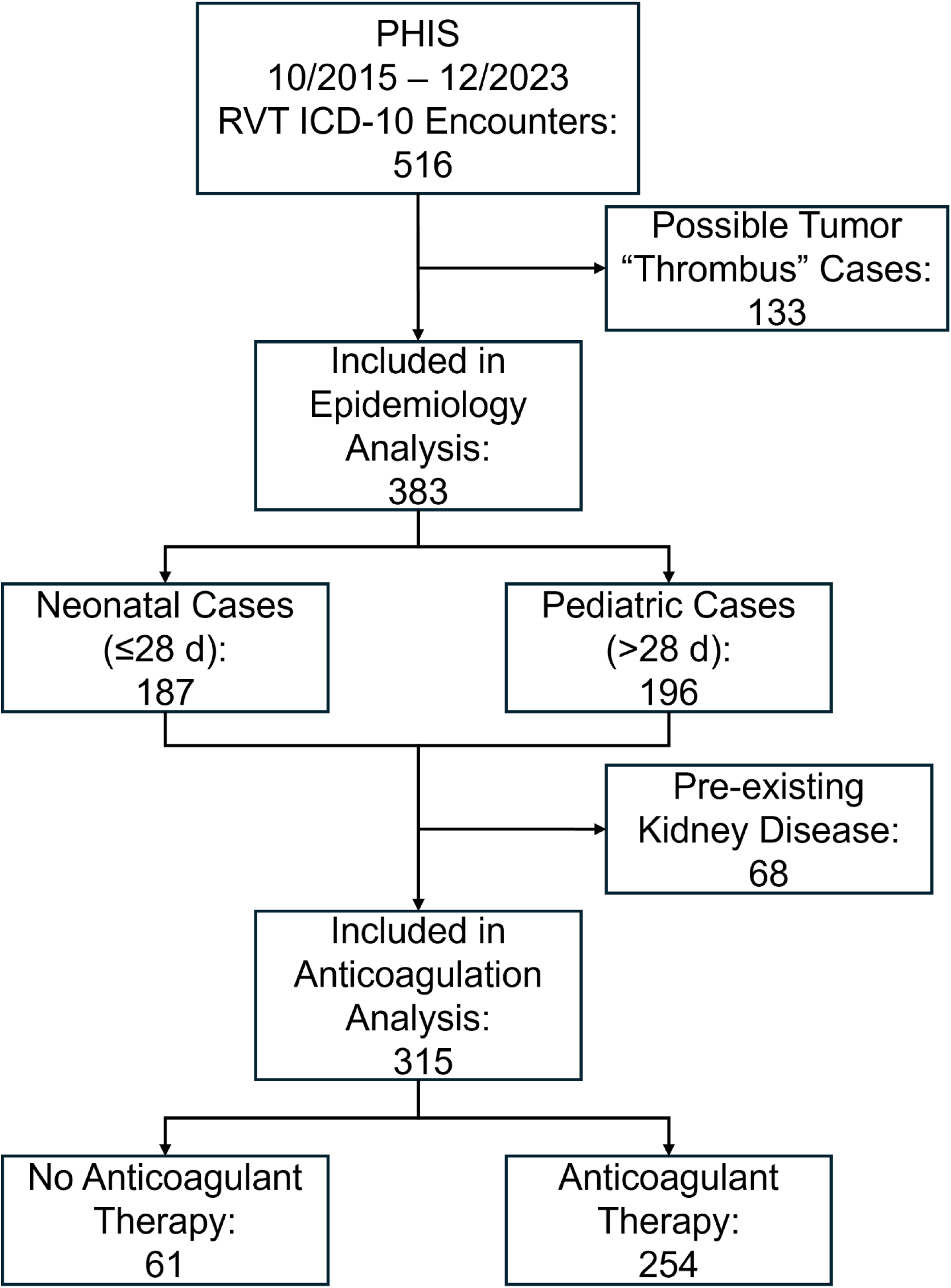
RVT Case Selection and Analytical Categorization. Possible tumor “thrombus” cases were eliminated to focus on true thrombi. RVT were analyzed separately for neonates (≤28 days old) and older children (>28 days – 18 years). RVT cases with pre-existing kidney disease were excluded form the analysis of potential renal protective effects of anticoagulant therapy.

Because the exact date of RVT diagnosis is not coded in PHIS, we approximated age at RVT as the age at admission for the hospital encounter during which RVT was first identified. Patients were stratified into neonatal (≤28 days at the time of RVT admission) and non-neonatal (>28 days at admission). Pharmaceutical billing codes were used to determine which RVT patients were treated with any of the following anticoagulants: enoxaparin, fondaparinux, heparin, warfarin, apixaban, rivaroxaban, or dabigatran (**Table S4**). Anticoagulant treatment was defined as anticoagulation within 7 days of RVT diagnosis. Because heparin is often used for non-treatment purposes (e.g., to prevent central venous catheter occlusion), heparin treatment was further defined to include resource utilization codes for daily therapeutic monitoring labs (i.e., activated partial thromboplastin time (332150) and/or heparin assay (334003)).

### Pre-RVT Co-Morbid Conditions

Co-morbid conditions are known to contribute to pediatric thrombosis risk.^11–13^ We thus, considered pre-RVT complex chronic condition diagnoses, as previously described, in our epidemiologic assessment of RVT risk.^11,12,14,15^ Moreover, because it is a well-known risk factor for RVT, we also assessed peri-RVT nephrotic syndrome (i.e. nephrotic syndrome diagnosed prior to or within 90 days after RVT diagnosis; **Table S5**).^16^

### Recurrent VTE

Because 5-10% of children with an incident VTE have recurrent thromboembolic disease, we assessed what proportion of the RVT cases had a preceding VTE.^17–20^ We also considered incident RVT as a risk factor for recurrent VTE in any other vascular territory.

### Statistics

All data were summarized using standard descriptive statistics. Frequency and percentage for qualitative variables and median and interquartile range (IQR) for quantitative variables. The prevalence of patients that developed CKD, hypertension, proteinuria, or hematuria following the RVT event were estimated with a percentage. Comparisons between renal outcomes between those that received and did not receive anticoagulation were performed using nonparametric statistics. When assessing renal outcomes, those with an identified diagnosis of CKD, hypertension, proteinuria, or hematuria prior to the diagnosis of RVT were excluded from the analysis. P-values were two sided and those <0.05 were considered statistically significant. Analyses were completed using SAS software, version 9.4 (SAS Institute, Cary, NC).

## RESULTS

### RVT Demographics and Co-Morbidities

During the study period, 516 children had an initial ICD-10 coded RVT event, of these 133 (25.8%) were excluded because of a coincident ICD-10 code indicating a cancer diagnosis that may be associated with tumor invasion into the renal vein and/or IVC (sometimes referred to as “tumor thrombus”; **Fig. 1**). Three hundred eighty-three (74.2%) RVT cases with 796 patient-years of available follow-up data were included in the epidemiologic analysis of RVT in children, 209 (54.6%) of which were male (**Table 1**). The median age at RVT diagnosis for the entire cohort was 0.1 years. Nearly half (187; 48.8%) of the children were neonates (≤28 d) at RVT diagnosis with a median age of 0 days, suggesting that many neonatal RVT were present at birth (**Fig. 2**). The median age of the non-neonatal RVT cases was 11.2 years. RVT had no significant racial or ethnic association. Sixty-eight (17.8%) of the RVT patients had pre-existing kidney disease with or without hypertension, all of whom were non-neonates. Twenty-five (6.5%) of the RVT were recurrent VTE (i.e., the patients had a preceding VTE in another anatomic location), again only in non-neonatal RVT cases. Ninety-seven (25.3%) RVTs occurred in children who also had a complex chronic condition, exclusive to non-neonatal cases. Thirty-five (9.1%) of the RVTs occurred after or within 90 days prior to a diagnosis of nephrotic syndrome, which has a known predilection for RVT; this phenomenon was seen in both neonates and older children.^16,21,22^ Fifty-eight (15.1%) of the RVT episodes resulted were associated with in-hospital death, but it is not possible to attribute mortality to RVT from these data.

**Figure 2:**
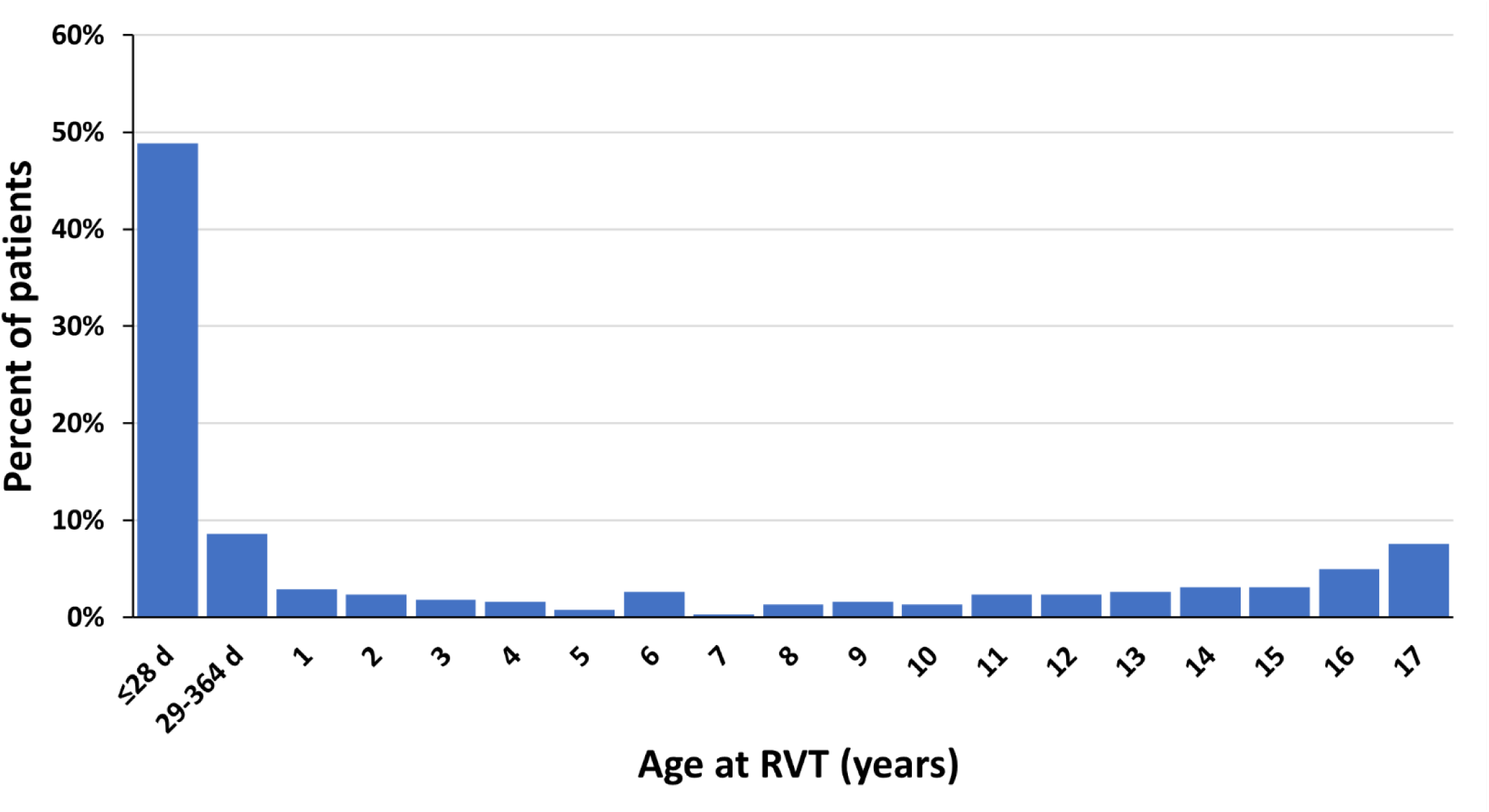
Age at RVT Diagnosis. Consistent with prior VTE data, age at RVT diagnosis was distributed in a bimodal fashion. Almost half (48.8%) of all RVT were diagnosed in neonates and another 51.2% in older children with a peak in late adolescence.

**Table 1:** Patient Characteristics.

| Characteristic | All Patients<br>*n* 383 | Age ≤28 days<br>*n* 187 | Age >28 days<br>*n* 196 | *p*-value<sup>1</sup> |
| --- | --- | --- | --- | --- |
| **Follow-up**; patient-years | 796 | 369 | 427 |  |
| **Post-RVT PHIS encounters**; median (IQR) | 5 (2-15) | 4 (2-13) | 7 (3-20) |  |
| **Male sex**; *n* (%) | 209 (54.6) | 113 (60.4) | 96 (49.0) | 0.024 |
| **Age at RVT**; median years (or days) (IQR) | 0.1 (0.01-11.4) | 0 (0-4) days | 11.2 (2.6-15.9) | - |
| **Race-ethnicity**; *n* (%) |  |  |  | 0.7 |
| Non-Hispanic White | 188 (49.1) | 98 (52.4) | 90 (45.9) |  |
| Non-Hispanic Black | 78 (20.4) | 38 (20.3) | 40 (20.4) |  |
| Hispanic/Latino | 72 (18.8) | 28 (15.0) | 44 (22.4) |  |
| Asian | 7 (1.8) | 4 (2.1) | 3 (1.5) |  |
| Other | 17 (4.4) | 8 (4.3) | 9 (4.6) |  |
| Multiracial | 9 (2.3) | 5 (2.7) | 4 (2.0) |  |
| Unknown | 12 (3.1) | 6 (3.2) | 6 (3.1) |  |
| **Pre-RVT kidney disease**<sup>2</sup>; *n* (%) | 68 (17.8) | 0 (-) | 68 (34.7) | < .0001 |
| Renal disease (CKD, AKI, infarction) | 56 (14.6) | 0 (-) | 56 (28.6) | < .0001 |
| Hypertension | 43 (11.3) | 0 (-) | 43 (21.4) | < .0001 |
| Proteinuria | 13 (3.4) | 0 (-) | 13 (6.6) | 0.0003 |
| Hematuria | 5 (1.3) | 0 (-) | 5 (1.3) | 0.06 |
| **Pre-RVT VTE diagnosis (non-RVT)**<sup>2</sup>; *n* (%) | 25 (6.5) | 0 (-) | 25 (12.8) | < .0001 |
| **Pre-RVT complex chronic conditions**<sup>2</sup>; *n* (%) | 97 (25.3) | 0 (-) | 97 (49.5) | < .0001 |
| Renal | 55 (14.4) | 0 (-) | 55 (28.1) | < .0001 |
| Cardiac | 35 (9.1) | 0 (-) | 35 (17.9) | < .0001 |
| Gastrointestinal | 44 (11.5) | 0 (-) | 44 (22.4) | < .0001 |
| Hematologic/immunologic | 26 (6.8) | 0 (-) | 26 (13.3) | < .0001 |
| Malignancy | 12 (3.1) | 0 (-) | 12 (6.1) | 0.0006 |
| Metabolic | 40 (10.4) | 0 (-) | 40 (20.4) | < .0001 |
| Neurologic | 16 (4.2) | 0 (-) | 16 (8.2) | < .0001 |
| Genetic/congenital | 15 (3.9) | 0 (-) | 15 (7.7) | 0.0001 |
| Respiratory | 10 (2.6) | 0 (-) | 10 (5.1) | 0.0018 |
| Technology dependent | 60 (15.7) | 0 (-) | 60 (30.6) | < .0001 |
| Neonatal | 12 (3.1) | 0 (-) | 12 (6.1) | 0.0006 |
| Transplant recipient | 20 (5.2) | 0 (-) | 20 (10.2) | < .0001 |
| **Peri-RVT nephrotic syndrome**<sup>3</sup>; *n* (%) | 35 (9.1) | 3 (1.6) | 32 (16.3) | < .0001 |
| **Mortality**; *n* (%) | 58 (15.1) | 37 (19.8) | 21 (10.7) | 0.0133 |

<sup>1</sup>Neonates (≤28 days) vs. Older Children (>28 days); <sup>2</sup>Defined as evidence of diagnosis prior to the first admission RVT diagnosis was observed;
<sup>3</sup>Defined as presence of nephrotic syndrome prior to RVT or within 90 days after initial RVT encounter. RVT: renal vein thrombosis; PHIS: pediatric health information system; VTE: venous thromboembolism

### Contemporaneous and Post-RVT Outcomes

Of the 383 patients with RVT, 164 (42.8%) developed clinically-relevant renal complications during or after their hospital stay (i.e., AKI, CKD, hypertension, and/or proteinuria; **Fig. 3**). Ninety-two (24%) patients had at least one AKI event, with 56 (60.9%) of those patients being in the non-neonatal group, and 36 (39.1%) in the neonatal group (**Table 2**). CKD developed in 85 (22.2%) of the children and was significantly more common in the non-neonatal group. Importantly, 41 (48.2%) of the children who developed CKD had ESKD which was also most likely amongst the non-neonatal RVT cases. Hypertension developed in 26.9% (103) of the RVT cases and was significantly more likely in the non-neonatal group. Proteinuria developed in 12 (3.1%) of the patients and was more common in older children. One hundred twenty-six (32.9%) RVT patients developed non-RVT venous thromboembolism during or after their hospital stay.

**Figure 3:**
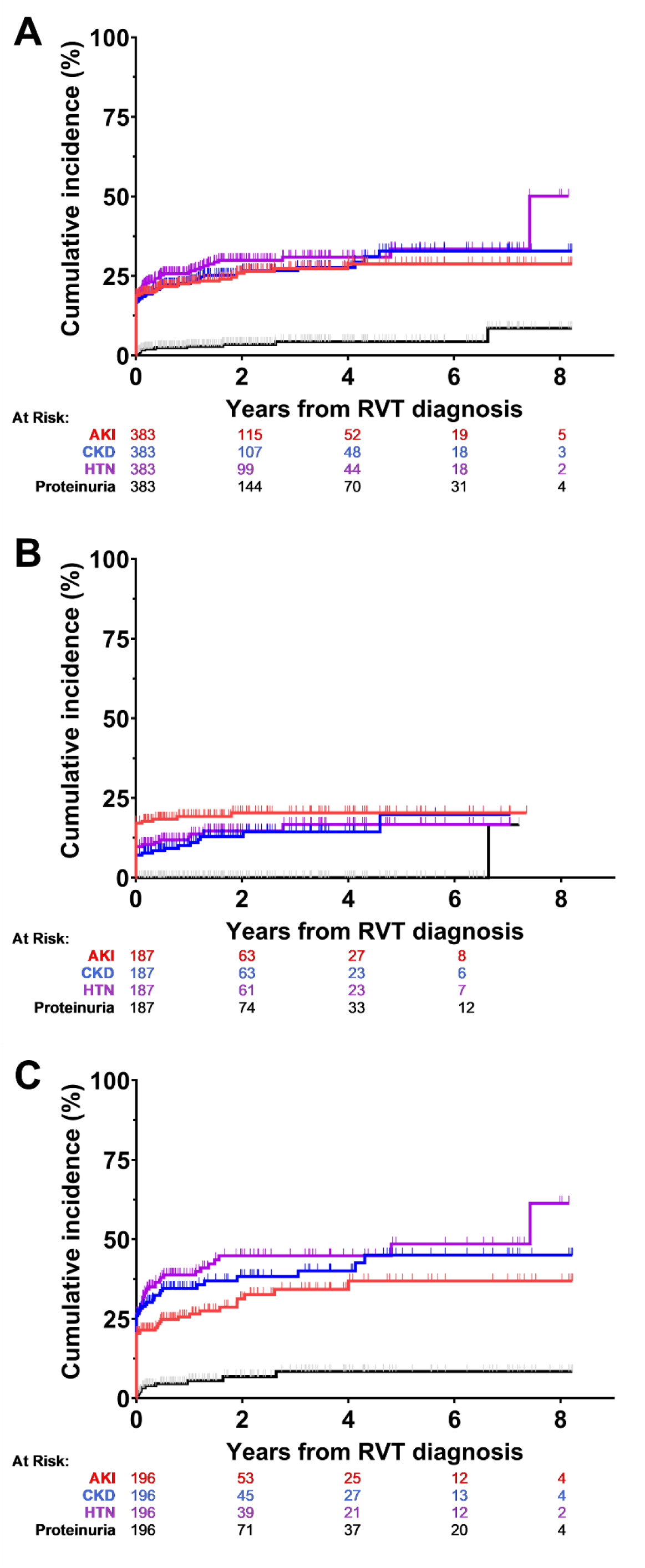
Most RVT-Related Kidney Disease Presents During the Same Admission that RVT is Diagnosed. Kidney disease outcomes for (A) all patients, (B) neonates, and (C) older children.

**Table 2:** RVT Outcomes of Interest.

| **Characteristic\*** | **All Patients**<br>***n* 383** | **Age ≤28 days**<br>***n* 187** | **Age >28 days**<br>***n* 196** | ***p*-value<sup>1</sup>** |
| --- | --- | --- | --- | --- |
| **Renal disease; *n* (%)** | 164 (42.8) | 59 (31.6) | 105 (53.6) | < .0001 |
| **AKI** | 92 (24.0) | 36 (19.3) | 56 (28.6) | 0.0328 |
| **Any CKD** | 85 (22.2) | 21 (11.2) | 64 (32.7) | < .0001 |
| CKD – stage I-IV | 35 (9.1) | 8 (4.3) | 27 (13.8) | 0.0013 |
| CKD – ESKD | 41 (10.7) | 6 (3.2) | 35 (17.9) | < .0001 |
| Dialysis Dependent | 23 (6.0) | 5 (2.7) | 18 (9.2) | 0.0074 |
| Kidney Transplant | 44 (11.5) | 2 (1.1) | 42 (21.4) | < .0001 |
| CKD – nonspecific | 30 (7.8) | 11 (5.9) | 19 (9.7) | 0.17 |
| Small Kidney/Atrophy | 19 (5.0) | 12 (6.4) | 7 (3.6) | 0.20 |
| Kidney Infarction | 19 (5.0) | 11 (5.9) | 8 (4.1) | 0.42 |
| **Hypertension** | 103 (26.9) | 25 (13.4) | 78 (39.8) | < .0001 |
| **Proteinuria** | 12 (3.1) | 1 (0.5) | 11 (5.6) | 0.0044 |
| **Hematuria** | 19 (5.0) | 11 (5.9) | 8 (4.1) | 0.42 |
| **Non-RVT VTE; *n* (%)** | 126 (32.9) | 57 (30.5) | 69 (35.2) | 0.33 |

\*Occurring during the same admission or anytime afterwards during follow-up. RVT: renal vein thrombosis; AKI: acute kidney injury; CKD: chronic kidney disease; ESKD: end-stage kidney disease; VTE: venous thromboembolism

### Treatment Patterns by Age Group

Of the 383 patients, 307 (80.2%) were treated with anticoagulation (**Table 3**). The proportion of children treated with anticoagulation did not differ by age group. With the exception of warfarin, which is difficult to manage in neonates, the choice of anticoagulant modality also did not appear to differ by age.^20,23^

**Table 3:**
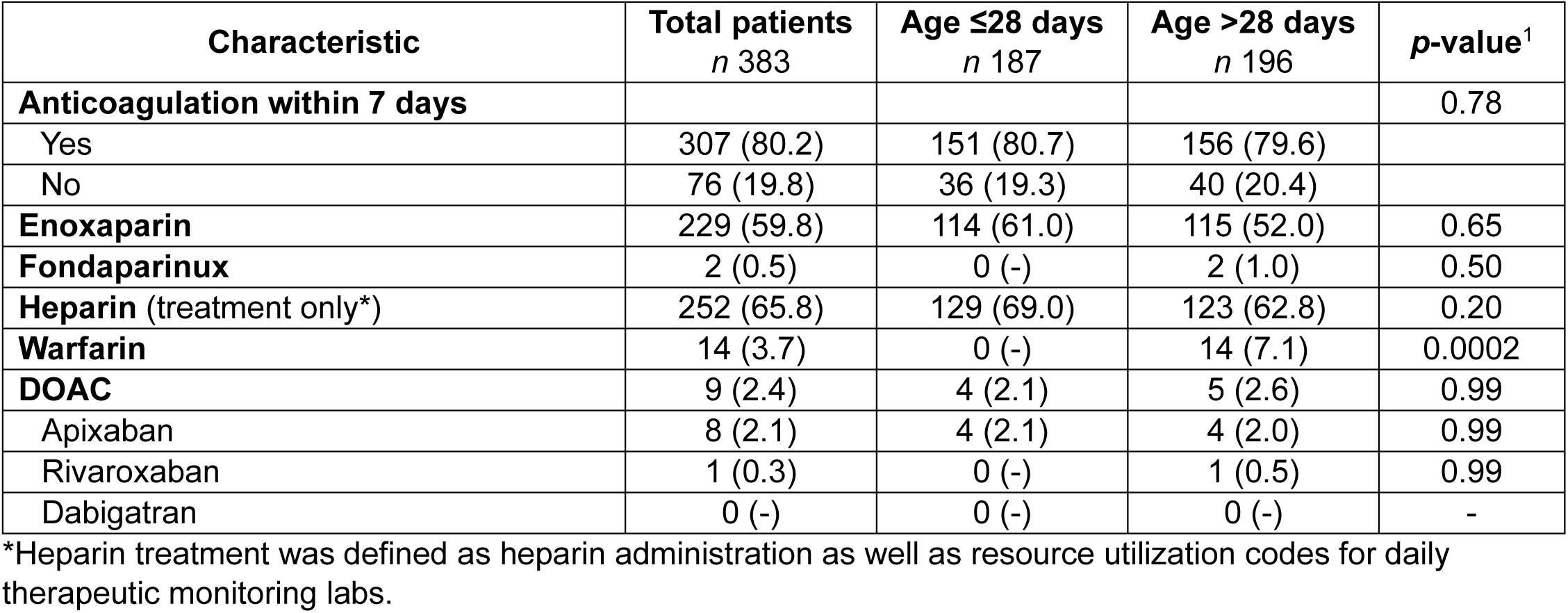
Anticoagulation Treatment.

| **Characteristic** | **Total patients**<br>***n* 383** | **Age ≤28 days**<br>***n* 187** | **Age >28 days**<br>***n* 196** | ***p*-value<sup>1</sup>** |
| --- | --- | --- | --- | --- |
| **Anticoagulation within 7 days** |  |  |  | 0.78 |
| Yes | 307 (80.2) | 151 (80.7) | 156 (79.6) |  |
| No | 76 (19.8) | 36 (19.3) | 40 (20.4) |  |
| **Enoxaparin** | 229 (59.8) | 114 (61.0) | 115 (52.0) | 0.65 |
| **Fondaparinux** | 2 (0.5) | 0 (-) | 2 (1.0) | 0.50 |
| **Heparin** (treatment only*) | 252 (65.8) | 129 (69.0) | 123 (62.8) | 0.20 |
| **Warfarin** | 14 (3.7) | 0 (-) | 14 (7.1) | 0.0002 |
| **DOAC** | 9 (2.4) | 4 (2.1) | 5 (2.6) | 0.99 |
| Apixaban | 8 (2.1) | 4 (2.1) | 4 (2.0) | 0.99 |
| Rivaroxaban | 1 (0.3) | 0 (-) | 1 (0.5) | 0.99 |
| Dabigatran | 0 (-) | 0 (-) | 0 (-) | - |

\*Heparin treatment was defined as heparin administration as well as resource utilization codes for daily therapeutic monitoring labs.

### Outcomes by Anticoagulant Treatment Group

In order to evaluate the potential effect of anticoagulation therapy on the likelihood of peri- or post-RVT kidney disease, 68 (17.7%) of the RVT patients who had pre-RVT renal disease, hypertension, proteinuria, or hematuria were excluded from analysis (**Table S3**). Of the remaining 315 patients, 254 (80.6%) were treated with anticoagulation (**Table 4**). Anticoagulation did not significantly alter the likelihood of RVT-related kidney disease overall or by age group. In contrast, anticoagulation was significantly associated with recurrent non-RVT venous thromboembolism.

**Table 4:** Outcomes of Interest by Anticoagulant Treatment Group.

|  | All Patients |  |  | Age ≤28 days |  |  | Age >28 days |  |  |
| --- | --- | --- | --- | --- | --- | --- | --- | --- | --- |
| **Post-RVT Outcomes\*** | **No AC**<br>**n 61** | **AC**<br>**n 254** | **p-value** | **No AC**<br>**n 36** | **AC**<br>**n 151** | **p-value** | **No AC**<br>**n 25** | **AC**<br>**n 103** | **p-value** |
| **Renal disease** | 17 (27.9) | 95 (37.4) | 0.16 | 8 (22.2) | 51 (33.8) | 0.18 | 9 (36.0) | 44 (42.7) | 0.54 |
| **AKI** | 11 (18.0) | 57 (22.4) | 0.45 | 6 (16.7) | 30 (19.9) | 0.66 | 5 (20.0) | 27 (26.2) | 0.52 |
| **Any CKD** | 7 (11.5) | 39 (15.4) | 0.44 | 4 (11.1) | 17 (11.3) | 0.99 | 3 (12.0) | 22 (21.4) | 0.40 |
| CKD – stage I-IV | 4 (6.6) | 14 (5.5) | 0.76 | 3 (8.3) | 5 (3.3) | 0.18 | 1 (4.0) | 9 (8.7) | 0.69 |
| CKD – stage V/ESKD | 2 (3.3) | 14 (5.5) | 0.75 | 1 (2.8) | 5 (3.3) | 0.99 | 1 (4.0) | 9 (8.7) | 0.69 |
| Dialysis dependent | 1 (1.6) | 8 (3.1) | 0.99 | 1 (2.8) | 4 (2.6) | 0.99 | 0 (-) | 4 (3.9) | 0.99 |
| Kidney Transplant | 2 (3.3) | 9 (3.5) | 0.99 | 1 (2.8) | 1 (0.7) | 0.35 | 1 (4.0) | 8 (7.8) | 0.99 |
| CKD – nonspecific | 6 (9.8) | 14 (5.5) | 0.24 | 4 (11.1) | 7 (4.6) | 0.23 | 2 (8.0) | 7 (6.8) | 0.99 |
| Small Kidney/Atrophy | 3 (4.9) | 15 (5.9) | 0.99 | 1 (2.8) | 11 (7.3) | 0.47 | 2 (8.0) | 4 (3.9) | 0.33 |
| Kidney Infarction | 1 (1.6) | 15 (5.9) | 0.33 | 0 (-) | 11 (7.3) | 0.13 | 1 (4.0) | 4 (3.9) | 0.99 |
| **Hypertension** | 13 (21.3) | 50 (19.7) | 0.78 | 5 (13.9) | 20 (13.2) | 0.99 | 8 (32.0) | 30 (29.1) | 0.78 |
| **Proteinuria** | 0 (-) | 6 (2.4) | 0.60 | 0 (-) | 1 (0.7) | 0.99 | 0 (-) | 5 (4.9) | 0.58 |
| **Hematuria** | 3 (4.9) | 13 (5.1) | 0.99 | 2 (5.6) | 9 (6.0) | 0.99 | 1 (4.0) | 3 (3.9) | 0.99 |
| **Non-RVT VTE** | 4 (6.6) | 99 (39.0) | < .0001 | 1 (2.8) | 56 (37.1) | < .0001 | 3 (12.0) | 43 (41.7) | 0.0054 |

\*Occurring during the same admission or any time after up to last follow-up encounter. RVT: renal vein thrombosis; AC: anticoagulation; AKI: acute kidney injury; CKD: chronic kidney disease; ESKD: end-stage kidney disease; VTE: venous thromboembolism

## DISCUSSION

In this large retrospective administrative database study, we found that AKI, CKD and hypertension are prevalent sequelae of childhood RVT. Importantly, CKD and hypertension are common complications of both neonatal and non-neonatal RVT. Remarkably, anticoagulant therapy had no effect on these important renal outcomes. In contrast to previous studies, we found that over half of RVT cases occurred in non-neonates.^2–5^ Pre-existing kidney disease, other complex chronic conditions, and prior VTE were all risk factors for development of RVT only in non-neonates. However, nephrotic syndrome was a risk factor for RVT in both age groups. Moreover, in children who did not have a prior VTE, RVT was associated with development non-RVT VTE. The majority of RVT were treated with anticoagulation despite the lack of high-quality evidence supporting its use.^7^ Counterintuitively, anticoagulant therapy for RVT was associated with an increased incidence of recurrent VTE.

Previous neonatal RVT outcome studies have documented CKD and hypertension as common complications and the risk for these outcomes persists for many years post-RVT.^4,24–26^ Our study also demonstrated that AKI, CKD, and hypertension occur with concerning frequency in children with RVT. Importantly, all three of these renal complications were significantly more common in older children than in neonates. This suggests that RVT carries previously unrecognized renal outcome risk for older children as well as neonates. Unfortunately, anticoagulation did not reduce the incidence of any of the renal outcomes examined in this study. However, extensiveness of RVT (e.g., bilateral RVT and/or extension into the vena cava) and other inciting factors (e.g., renal mass, catheter, etc.) are known to influence the decision to prescribe anticoagulation.^1,7,25^ Regrettably, it is not possible to discern RVT extent or AKI severity using administrative data, both of which are likely to have impacted both the decision to treat and the likelihood of future CKD and hypertension. While severe AKI increases the risk of CKD independent of anticoagulation, it is also possible that anticoagulation may facilitate renal recovery and lessen the likelihood of progression to CKD. It is not possible to analyze these complex relationships from the available administrative data. It is also reasonable to postulate that RVT extent may have an influence on mortality.

In contrast to previous literature suggesting that the majority of pediatric RVT occurs in neonates, we found a relatively equal distribution of RVT between neonates and older children.^2–5^ However, the age distribution of RVT in this study is consistent with the well-known bimodal age distribution for pediatric VTE in general.^11–13,27,28^ Known neonatal RVT risk factors are prematurity, birth hypoxia, sepsis, dehydration, maternal diabetes, and thrombophilia.^1,6,25,26^ Other than the well-known association with nephrotic syndrome, RVT risk factors for older children are ill-defined.^16,21,22^ Here we show that complex chronic conditions of essentially any organ system increase RVT risk. Meanwhile the influence of thrombophilia on RVT risk in older children is not well understood. However, previous non-RVT VTE was identified as an RVT risk factor and may reflect recurrent thromboembolic disease due to an underlying thrombophilia. Meanwhile, VTE at another site developed in about ⅓^rd^ of the RVT cases and was significantly more common in the patients who received therapeutic anticoagulation. This counterintuitive observation may be due to confounding by indication wherein the providers chose to prescribe anticoagulation for children perceived to be most ill and thus at highest risk for multifocal and/or recurrent VTE. Moreover, the fact that most children (∼80%) were anticoagulated may indicate a bias towards treatment, despite the low-quality evidence supporting its use.^7^ While a randomized controlled trial would facilitate a better understanding of the role of anticoagulation in pediatric RVT management it is unlikely that a such an effort would succeed given the rarity of RVT.

While administrative data are a valuable resource to enable the study of rare diseases, such as RVT, they come with several limitations. Overall data accuracy depends on ICD-10 coding precision, which is subject to both miscoding and non-coding. While ICD codes for have demonstrated high positive predictive values in adults, a small study suggests that they are less reliable for pediatric VTE.^29,30^ Meanwhile, the sensitivity and specificity of ICD codes for the other pediatric conditions included in the present study are relatively unknown. Nonetheless, the use of ICD codes for the study of pediatric VTE and complex chronic conditions using administrative data has proven to be a well-established and valuable method.^11–15,28,31^ An inherent problem is the inability to independently validate RVT and other VTE ICD-10 codes through examination of imaging data. Central venous catheters are a major risk factor for pediatric VTE but the presence or absence of these devices is difficult to ascertain from administrative datasets.^32–36^ The study of a larger timeframe may have enhanced power, but ICD-10 replaced ICD-9-CM coding in 2015 which would have made it difficult to obtain consistent data via extension to an earlier data collection start date.

This administrative data study demonstrated that RVT occurs equivalently in both neonates and older children. AKI, CKD, and hypertension frequently occur during the same admission as RVT diagnosis, but their incidence continues to rise for several years afterwards. Therefore, survivors of childhood RVT should receive kidney health assessments on a regular basis. Unfortunately, anticoagulation therapy does not seem to reduce the likelihood of adverse renal outcomes. Further studies comparing anticoagulation therapy among patients stratified for RVT extent are needed to better define the role of anticoagulation in the prevention of renal sequelae. In addition to active monitoring for renal complications, RVT survivors should receive prophylactic anticoagulation during high-risk episodes (e.g., major surgery, hospitalization) due to their elevated risk for recurrent VTE.

## Data Availability

All data produced in the present work are contained in the manuscript

## ACKNOWLEDGEMENTS

Portions of this work were presented as an abstract at the annual meeting of the American Society of Nephrology.

## Author contributions

MAO, LSB, JRS, MK, and BAK developed the data extraction strategy. MAO, LSB, and JRS analyzed the data. JRS performed statistics, tabulated data, and drafted the figures. MAO and BAK wrote the paper. MAO, MK, and BAK conceived the study. BAK provided financial support. All authors refined the final draft.

