## Supplementary material for "Renal Outcomes in Survivors of Neonatal and Pediatric Renal Vein Thrombosis": Table S1

**Table S1: RVT and VTE ICD-10 codes**

| Diagnosis | ICD-10 Codes |
|---|---|
| RVT | I82.3 |
| All other VTE | I81, I82.0, I82.220, I82.221, G08, I80.3, I82.401, I82.402, I82.403, I82.409, I82.411, I82.412, I82.413, I82.419 I82.421, I82.422, I82.423, I82.429, I82.431, I82.432, I82.433, I82.439, I82.441, I82.442, I82.443, I82.449, I82.491, I82.492, I82.493, I82.499, I82.4Y1, I82.4Y2, I82.4Y3, I82.4Y9, I82.4Z1, I82.4Z2, I82.4Z3, I82.4Z9, I82.501, I82.502, I82.503, I82.509, I82.511, I82.512, I82.513, I82.519, I82.521, I82.522, I82.523, I82.529, I82.531, I82.532, I82.533, I82.539, I82.541, I82.542, I82.543, I82.549, I82.591, I82.592, I82.593, I82.599, I82.5Y1, I82.5Y2, I82.5Y3, I82.5Y9, I82.5Z1, I82.5Z2, I82.5Z3, I82.5Z9, I80.8, I80.9, I82.1, I82.890, I82.891, I82.90, I82.91, I26.02, I26.09, I26.92, I26.99, I27.82, I51.3, I82.210, I82.211, I82.290, I82.291, I82.601, I82.602, I82.603, I82.609, I82.611, I82.612, I82.613, I82.619, I82.621, I82.622, I82.623, I82.629, I82.701, I82.702, I82.703, I82.709, I82.711, I82.712, I82.713, I82.719, I82.721, I82.722, I82.723, I82.729, I82.A11, I82.A12, I82.A13, I82.A19, I82.A21, I82.A22, I82.A23, I82.A29, I82.B11, I82.B12, I82.B13, I82.B19, I82.B21, I82.B22, I82.B23, I82.B29, I82.C11, I82.C12, I82.C13, I82.C19, I82.C21, I82.C22, I82.C23, I82.C29 |

RVT: Renal Vein Thrombosis; VTE: Venous thromboembolism
