## Supplementary material for "Renal Outcomes in Survivors of Neonatal and Pediatric Renal Vein Thrombosis": Table S2

**Table S2: Cancers potentially associated with tumor "thrombus"**

| Diagnosis | ICD-10 Codes |
|---|---|
| Cancers or tumors of the pelvis or kidney (e.g., Wilms tumor, NBL) | C48.0, C48.8, C49.4, C62.11, C62.12, C62.90, C62.91, C62.92, C64.1, C64.2, C64.9, C65.1, C65.2, C65.9, C74.00, C74.01, C74.02, C74.90, C74.91, C74.92, C76.2, C78.6, C79.00, C79.01, C79.02, C79.70, C79.71, C79.72, D17.71, D41.00, D41.01, D41.02, D49.511, D49.512, D49.519 |

NBL: Neuroblastoma
