## Supplementary material for "Renal Outcomes in Survivors of Neonatal and Pediatric Renal Vein Thrombosis": Table S3

**Table S3: Kidney disease ICD-10 codes**

| Diagnosis | ICD-10 Codes |
|---|---|
| AKI | N17.0, N17.1, N17.2, N17.8, N17.9 |
| CKD-Stage I-IV | N18.1, N18.2, N18.3, N18.30, N18.31, N18.32, N18.4 |
| CKD Stage V or ESKD | N18.5, N18.6, N19 |
| CKD non-specific stage | N18.9 |
| Hypertension | I10, I12.0, I12.9, I15.0, I151, I15.8, I15.9, I1A.0, I13.0, I13.10, I13.11, I13.2, R03.0, P29.2 |
| Proteinuria | R80.0, R80.1, R80.8, R80.9 |
| Hematuria | R31.0, R31.2, R31.29, R31.9 |

AKI: Acute kidney injury; CKD: Chronic kidney disease; ESKD: End-stage kidney disease
