## Supplementary material for "Renal Outcomes in Survivors of Neonatal and Pediatric Renal Vein Thrombosis": Table S4

**Table S4: Anticoagulant pharmaceutical billing codes**

| Medications | PHIS Clinical Transaction Codes (CTC) |
|---|---|
| Enoxaparin | 161327 |
| Fondaparinux | 161341 |
| Heparin | 161307 |
| Warfarin | 161361 |
| Apixaban | 161352 |
| Rivaroxaban | 161356 |
| Dabigatran | 161354 |

PHIS: Pediatric Health Information System database
