## Supplementary material for "Renal Outcomes in Survivors of Neonatal and Pediatric Renal Vein Thrombosis": Table S5

**Table S5: Nephrotic Syndrome ICD-10 codes**

| Diagnosis | ICD-10 Codes |
|---|---|
| Nephrotic syndrome | N00.0, N00.1, N00.2, N00.3, N00.4, N00.5, N00.6, N00.7, N00.8, N00.9, N00.A, N01.0, N01.1, N01.2, N01.3, N01.4, N01.5, N01.6, N01.7, N01.8, N01.9, N01.A, N02.0, N02.1, N02.2, N02.3, N02.4, N02.5, N02.6, N02.7, N02.8, N02.9, N02.A, N02.B1, N02.B2, N02.B3, N02.B4, N02.B5, N02.B6, N02.B9, N03.0, N03.1, N03.2, N03.3, N03.4, N03.5, N03.6, N03.7, N03.8, N03.9, N03.A, N04.0, N04.1, N04.2, N04.20, N04.21, N04.22, N04.29, N04.3, N04.4, N04.5, N04.6, N04.7, N04.8, N04.9, N04.A, N05.0, N05.1, N05.2, N05.3, N05.4, N05.5, N05.6, N05.7, N05.8, N05.9, N05.A |
